# When Dose Matters: Linking Exposure, Environment, and Epidemic Persistence in Wildlife Systems

**DOI:** 10.64898/2026.03.10.26348022

**Authors:** Mohammad Mihrab Chowdhury, E. Davis Carter, Matthew J. Gray, Angela Peace

## Abstract

Understanding how infectious disease spreads through exposure is key to predicting outbreaks and effective control measures. This is important for wildlife, where outbreaks often lead to devastating ecological consequences. Traditional epidemiological models often assume equal infection risk regardless of exposure dose and focus on a single infection stage. This assumption eventually overlook variations in initial doses and the role of environmental pathogen reservoirs. We hypothesized that higher exposure doses accelerate disease progression, increase mortality, and elevate pathogen shedding but reduce infectious periods. To evaluate this, we estimated key disease parameters - including latency period, disease-induced mortality, zoospore shedding rates, and transmission probability-from datasets of eastern newt (*Notophthalmus viridescens* ) exposed to varying zoospore concentrations of fungal pathogen *Batrachochytrium salamandrivorans (Bsal)*. Guided by the empirical evidence,we developed a novel mathematical model capturing dose-dependent transmission, multiple exposure stages, and environmental pathogen accumulation. This empirically driven modeling framework reveals how exposure levels shape disease trajectory, host outcomes and pathogen persistence. Our findings indicate that high initial doses lead to faster disease progression and higher mortality. On the other hand, environmental transmission can sustain outbreaks even under low direct contact rates. Integrating dose dependence with environmental transmission, our framework advances epidemiological modeling of pathogen persistence.

## 1. Introduction

Epidemiological studies have long emphasized the importance of transmission dynamics in understanding pathogen spread and persistence. Early disease models primarily focused on host-pathogen interactions within a homogeneously mixed population (Anderson and May, 1991). However, recent research highlights the role of exposure dosage in infectious disease dynamics (Li and Handel, 2014). The concept of exposure-dependent transmission is particularly relevant in viral and fungal epidemics, including flu-like diseases in humans (COVID-19, MERS) (Pujadas et al., 2020; Dadras et al., 2022; Cevik et al., 2021; Feikin et al., 2015; Lee et al., 2009) and epizootics in wildlife, such as chytrid fungus and ranavirus in amphibians (Brunner et al., 2005; Stockwell et al., 2010; Wu, 2023). Unlike human diseases, where medical interventions mitigate disease spread, wildlife diseases often progress unchecked (Keesing et al., 2010; Smith et al., 2009; Pedersen et al., 2007). This leads to devastating ecological consequences and species extinctions (Skerratt et al., 2007). This highlights the need for modeling approaches to capture the complexity of exposure-dependent disease dynamics across biological systems.

Traditional epidemiological models, such as the Susceptible-Infectious-Recovered (SIR) framework, often assume uniform infection probabilities across individuals and do not account for exposure dose. This overlook brings limitation in understanding epidemic progression and intervention strategies such as dose-dependent immune responses, incubation periods, and differential mortality rates. However, recent experimental evidence suggests that transmission probabilities vary significantly depending on the pathogen load (Li and Handel, 2014; Carter et al., 2021; Gray et al., 2023). Specifically, in fungal diseases, hosts can be infected through multiple routes: direct contact with an infected individual or exposure to zoospores in the environment (Chowdhury et al., 2025; Carter et al., 2021; Scheele et al., 2019). This dual transmission pathway creates a complex landscape in which host susceptibility varies over exposure to the environmental zoospore pool. Exposure dosage alters disease progression, affecting infection duration and survival time (Carter et al., 2021; Brunner et al., 2005; James et al., 2015).

To address these gaps, we developed a novel mathematical model using ordinary differential equations (ODEs) to capture the exposure-dependent dynamics. Our framework incorporates multiple exposure levels to explore the impact of variations in pathogen load. We introduce a dose-dependent transmission function in which infection risk varies with environmental pathogen concentration. The model differentiates individuals disease progression pathways depending on their exposure dose. Additionally, we included a zoospore compartment to accommodate dynamic changes in pathogen levels and their impact on environmental transmission. This is particularly important for diseases with environmental transmission, such as fungal and bacterial infections in wildlife and plants. Our model explicitly accounts for both direct (host-to-host contact) and environmental transmission (zoospore- or vector-mediated infection), reflecting real-world dynamics. Lastly, our model simulates interventions like pathogen-consuming micro-predators, providing actionable insights for disease control.

To demonstrate our model’s applicability, we analyze *Bsal* transmission dynamics on the eastern newt (*Notophthalmus viridescens* ). Salamanders are ecologically significant species. They contribute to ecosystem stability by regulating insect abundances and nutrient cycling and serving as a prey for predators (Hocking and Babbitt, 2014; Galatowitsch et al., 1999; Morin, 1985; Walters, 1975; Petranka et al., 1987; Walters, 1975).There are approximately 190 documented species of salamanders in USA (Institute, Accessed June 19, 2023), and the eastern newt is one of the most widely distributed species. On the other hand, *Bsal* has been solely responsible for declination of several salamander species in Europe and yet to emerge in USA (Martel et al., 2013; Laking et al., 2017; Spitzen-van der Sluijs et al., 2016; Sabino-Pinto et al., 2015; Cunningham et al., 2015; Thien et al., 2013). However, in laboratory experiments, it has proven lethal to U.S. salamander species(Richgels et al., 2016; Gray et al., 2015; Grisnik et al., 2023; Gray et al., 2023). Given its broad distribution, high susceptibility, and multiple transmission pathways, the eastern newt–Bsal system provides an ideal framework for applying our model (Carter et al., 2024; Petranka, 1998).

To quantify the effects of dose-dependent transmission, we conducted controlled laboratory experiments at the University of Tennessee, Knoxville. Eastern newts were exposed to a range of *Bsal* concentrations. Individuals were monitored following exposure, and data were collected on infection progression, infection loads, and host mortality (Carter et al., 2021). As a possible control strategy, we also collected data on zoospore predation by micro-predator Appendix E. These experimentally derived measurements were used to parameterize the model, and simulate the dose dependent disease dynamics.

Our model advances epidemiological methods by integrating exposure-dependent disease dynamics with environmental pathogen reservoirs. This integration creates a more biologically realistic representation of how infections develop and persist. The framework is flexible and can be applied to systems where dose, exposure routes, and environmental persistence shape transmission. Its application to *Bsal* illustrates its value in amphibian disease ecology and highlights its potential for studying other exposure-dependent pathogens.

## 2. Modeling Framework

Our framework explicitly accounts for dose-dependent transmission, multiple exposed and infectious stages, and environmental pathogen persistence. Unlike traditional models that assume uniform infection probabilities, our approach incorporates variations in exposure doses. By structuring the framework with multiple compartments, we track how the accumulation of environmental pathogen influences direct and indirect transmission. Building on this framework, we developed three compartmental models to capture the impact of exposure dose on disease dynamics. Each model represents a stepwise refinement of the classical SEIR epidemic framework. Our final model explicitly accounts for incorporating dose-response effects and environmental reservoirs.

The first model (Figure 1a stages of infection) extends the classical SEIR framework by introducing multiple infectious compartments. This approach is based on (Lloyd, 2001), which showed that dividing the infectious class into multiple stages approximates a more realistic, gamma-distributed infectious period. This modification alters the dynamic of outbreaks without changing the overall transmission structure. It serves as a foundation for representing heterogeneity in infectiousness and survival time. The second model (Figure 1b stage-driven transmission) incorporates exposure heterogeneity by dividing exposed and infectious compartments into multiple classes based on the initial exposure. Here, individuals follow different disease progression pathways depending on exposure level. For COVID19, Cheng et al. (2020) and Goyal et al. (2020) showed that infection risk varies over the course of the infectious period. Similar timing-dependent transmission patterns have been reported for norovirus (Zelner et al., 2013). However, these model exclude environmental transmission, assuming that exposure occurs via contact with infectious individuals only. Our final model (Figure 1c environmental load-driven transmission) incorporates exposure-dependent transmission and an explicit environmental zoospore reservoir. In addition to multiple pathways for direct transmission, we include routes for environmental transmission driven by the concentration of zoospores in the environment. As the zoospore pool fluctuates, the environmental transmission dynamics also change. We track stage-specific shedding rates across the infectious classes, as well as the resulting accumulation or decline of zoospores in the environment. This framework captures the dynamic relationship between zoospore shedding, environmental zoospore levels, and their combined influence on environmental transmission and overall disease dynamics.

**Figure 1.**
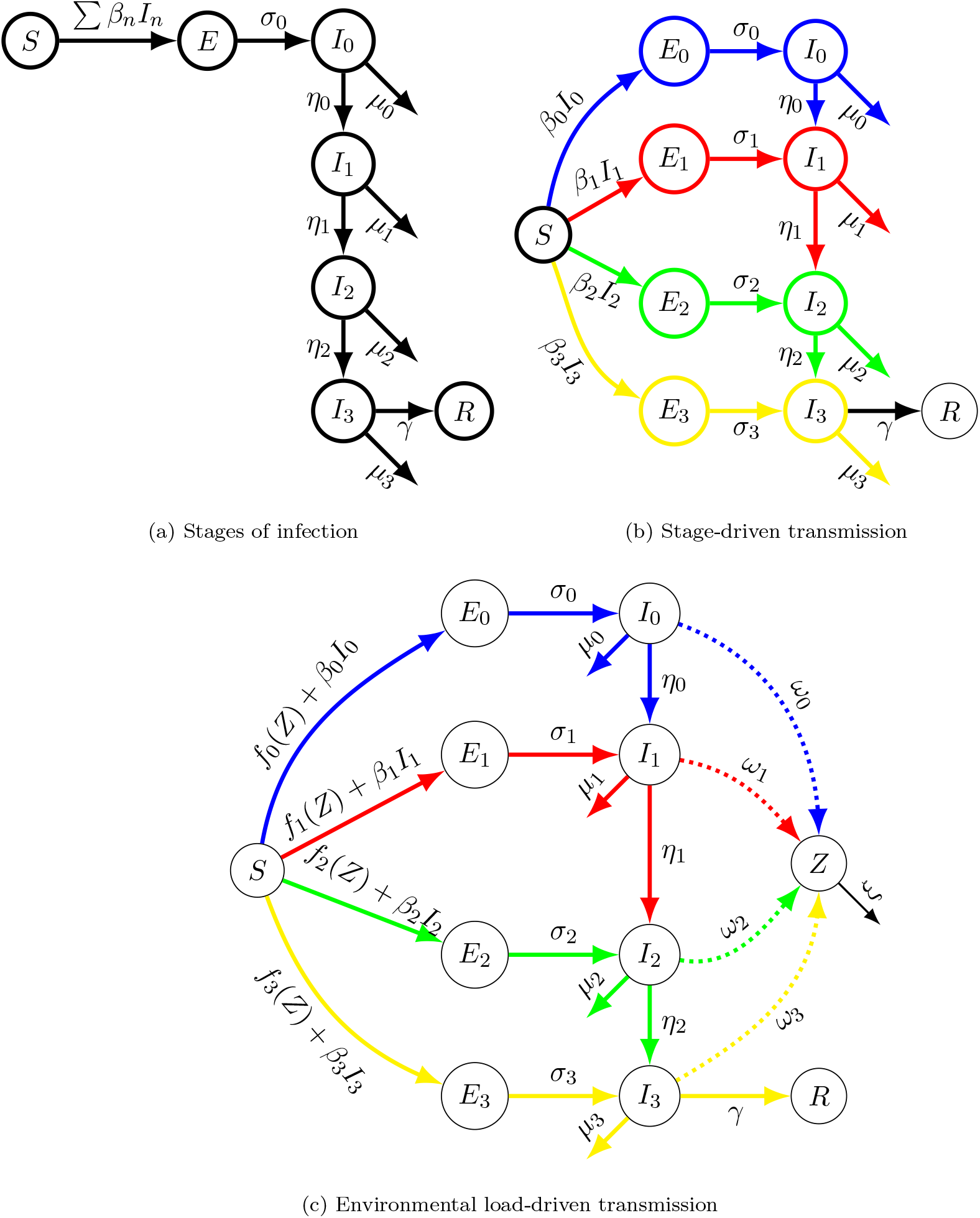
Comparative Schematic of Exposure-Dependent Disease Dynamics Models Incorporating DoseResponse and Environmental Transmission

## 3. Application

### 3.1. Eastern Newt and Basl

We applied (Figure 1c environmental load-driven transmission) to capture the biological and ecological processes underlying *Bsal* –eastern newt dynamics. The model captures both direct transmission and environmental exposure to zoospore through multiple exposed and infectious stages. Based on empirical evidence, we subsequently revised the direct-transmission pathway (Figure 2 *Bsal* - eastern newt model). Because eastern newts engage in only brief physical contact (Malagon et al., 2020), direct transmission is assumed to move individuals only into the initial exposed class (*E*_0_). However, varying levels of environmental zoospore results in additional exposed and infectious classes.

**Figure 2.**
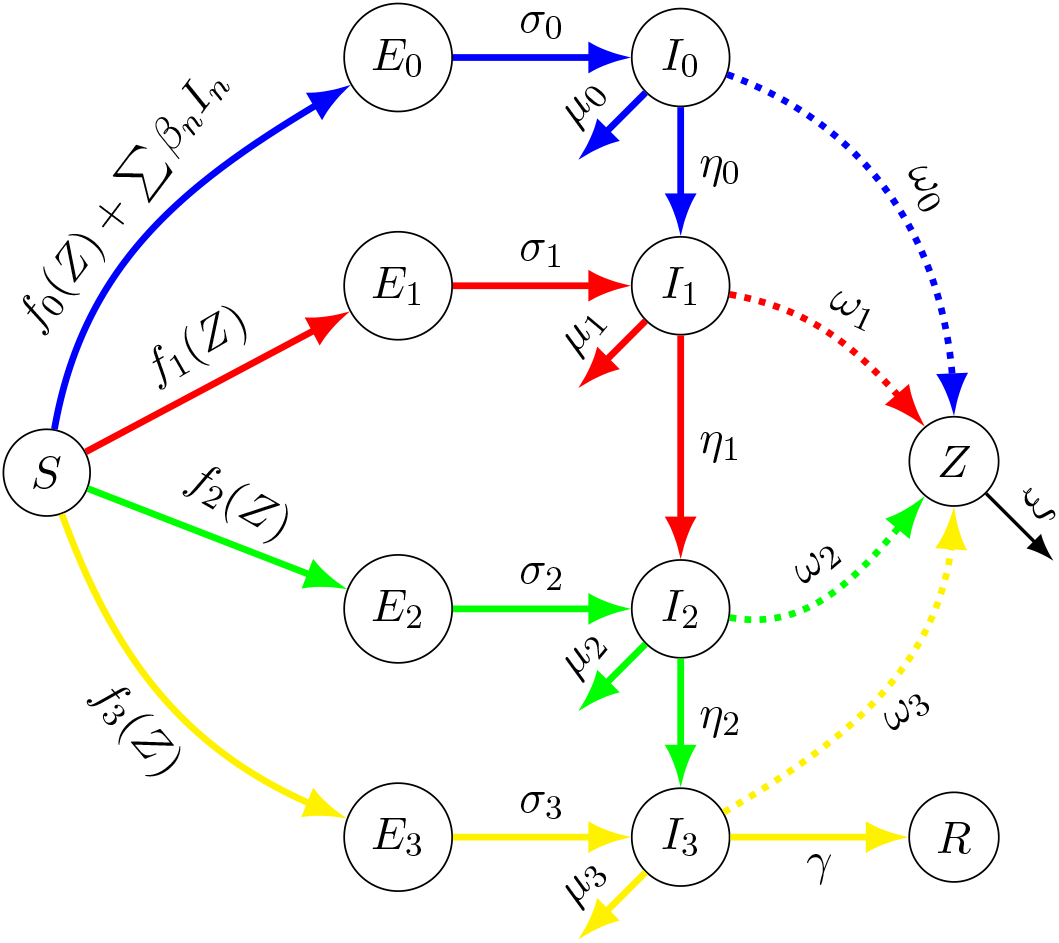
*Bsal* - eastern newt model

For our framework, we specifically focus on the dynamics between adult eastern newt and *Bsal*. Eastern newts have three life stages: larvae, eft, and adult. While efts live in terrestrial environments, larvae and adults reside in confined aquatic habitats. On the other hand, *Batrachochytrium salamandrivorans (Bsal)*, which means “salamander-eating chytrid fungus”, infects hosts by consuming keratin, making susceptibility stage-dependent. Larvae, lacking keratin except in their mouths, are largely resistant to infection (Petranka, 1998; Frost et al., 2006). In contrast, efts and adults have keratin on their skin and are highly susceptible to *Bsal* (Shepack and Catenazzi, 2020). Adults remain in confined habitats, resulting in higher contact rates and increased exposure to environmental aquatic zoospores. This makes them ideal for our framework, which emphasizes dose-dependent and environment-driven transmission.

### 3.2. Model Compartments

Susceptible newts are exposed to *Bsal* through two primary routes: direct contact with infected individuals and environmental exposure to zoospores. The probability of transmission dynamically depends on the infection stage of the host and the exposure to zoospore dosage. To capture these, our model consists of one susceptible (*S*) compartment, four exposed (*E*_0_, *E*_1_, *E*_2_, *E*_3_) compartments, four infected (*I*_0_, *I*_1_, *I*_2_, *I*_3_) compartments, and one recovered (*R*) compartment. Although reinfection is not included in the current model, the *R* compartment allows its future integration. We also introduce a zoospore (*Z*) compartment to monitor environmental pathogen accumulation. These compartments facilitate the study of disease progression, spillover, and population dynamics under different exposure conditions. The schematic diagram (Figure 2 *Bsal* eastern newt model model ) illustrates our proposed framework for tracking disease dynamics based on initial exposure levels for eastern newt-*Bsal*.

### 3.3. Direct Transmission

To accurately characterize direct transmission, we considered contact rates and the probability of disease transmission upon contact. Empirical evidence from (Chowdhury et al., 2025) suggests that adult contact rates are density-dependent. We assumed the same density-dependent Holling Type-2 function from our previous paper (Chowdhury et al., 2025).

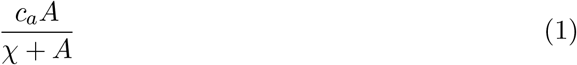

Where *c*_*a*_ is the maximum contact rate, *χ* is the half-saturation constant, and *A* is the density of the adult population. Moreover, due to the variability of the zoospore on the skin, we modeled the probability of transmission upon contact as host stage dependent. To account for this, our model assigns a unique direct transmission rate (*β*_*i*_ for *i* = 0 ™ 3) to each infectious stage. However, direct transmission has been observed with less than one second of contact (Malagon et al., 2020), and adults contact each other briefly. Given this rapid transmission, we assume all newly infected salamanders enter the *E*_0_ class regardless of the exposure.

### 3.4. Environmental Transmission

For the environmental transmission dynamics, we incorporated both zoospore persistence and host exposure. The environmental zoospore pool is driven by the shedding rate of infected newts, which changes as the infection progresses. To represent this, we assigned stage-specific shedding rates, *ω*_0_, *ω*_1_, *ω*_2_, and *ω*_3_, corresponding to each infected class. We assumed that baseline probability of contact between environmental zoospores and adult newts, denoted *b*_*a*_ is constant (Islam et al., 2021). Successful transmission depends on the probability of zoospore encystment on the skin, *ρ*. We also parameterized the zoospore dose required to infect 50% of susceptible hosts, *κ*, from the literature (Carter et al., 2021), and included the zoospore degradation rate, *ξ*. From our laboratory experiments detailed in ((Carter et al., 2021; Chowdhury et al., 2025), we obtained thresholds for the environmental zoospore concentration, *Z*. By exposing newts to a range of zoospore concentrations and measuring their subsequent shedding rates ((Carter et al., 2021)), we found that shedding levels depend on the initial exposure dose. This revealed an exposure-dependent threshold associated with the environmental zoospore concentration, *Z*.

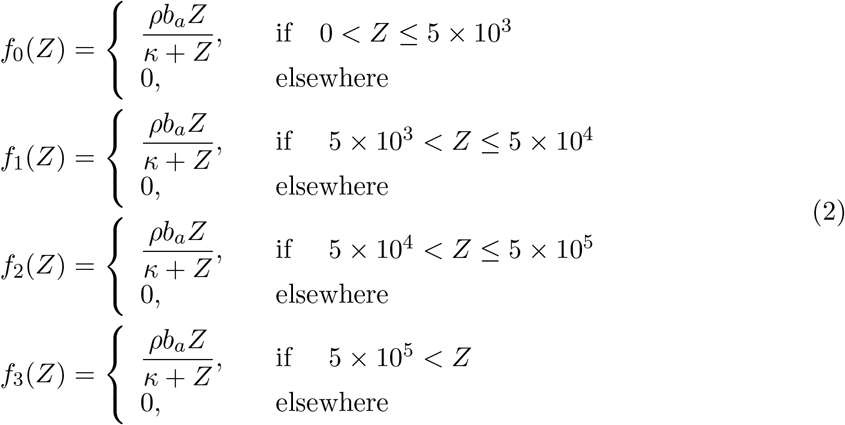

As the zoospore concentration fluctuates, the environmental transmission shifts up or down depending on the threshold in Equation 2.

We have incorporated the recovery rate, (*γ*), to account for the potential recovery of adults. Specifically, we did not consider recovery from distinct *I* classes. In our model, *I* classes represent disease progression over time. But our model can be expanded to include different recovery rates if the individual chance of recovery varies over time. However, as the infected individual progresses through the disease, the disease-induced mortality increases (*δ*_0_, *δ*_1_, *δ*_2_, *δ*_3_).

### 3.5. Model Equations

Taking all these things into account, the initial exposure-dependent disease dynamics is below:

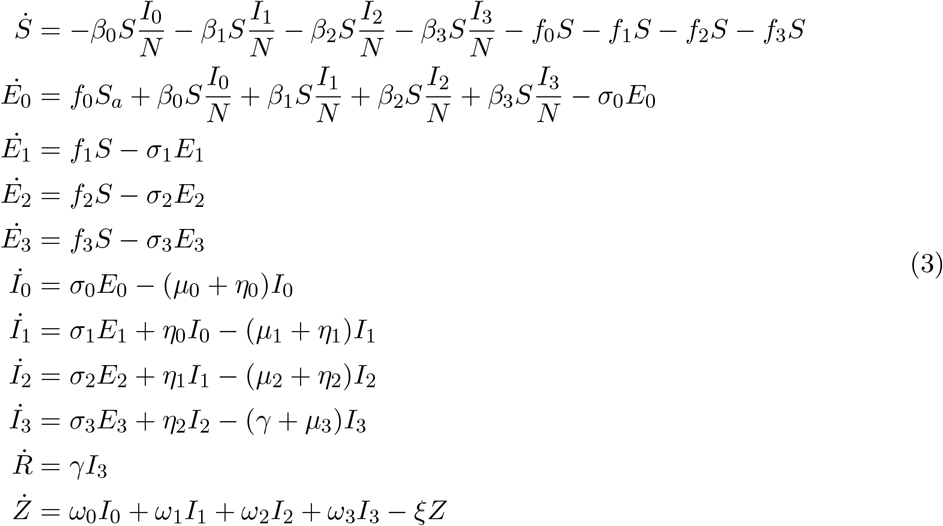

## 4. Model Parameterization

We estimated and identified the parameters of our model (3) by analyzing data from laboratory experiments and the literature. Model parameters are listed in Table 2.

### 4.1. Experimental Data Analysis

#### Initial Exposure Dosage

From the published data in (Carter et al., 2021), we found fluctuations in the disease dynamics depending on the exposure to the zoospore concentration. Varying zoospore concentration leads to multiple classes of exposed which in turn changes the incubation period (*σ*_0_, *σ*_1_, *σ*_2_), shedding rate (*ω*_0_, *ω*_1_, *ω*_2_, *ω*_3_), and disease-induced death rate (*µ*_0_, *µ*_1_, *µ*_2_, *µ*_3_) (Table 1).

**Table 1.**
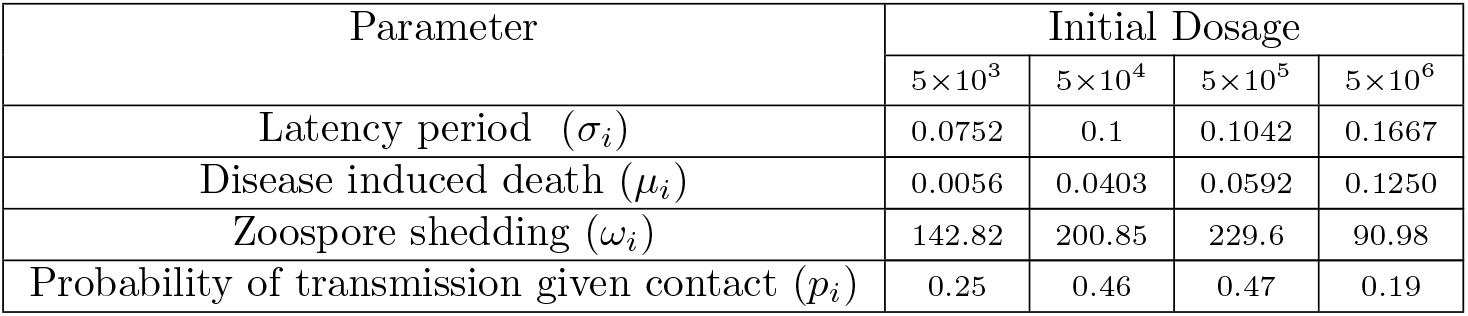
Impact of Initial Exposure Dosage (Carter et al., 2021; Gray and Carter, 2022-2024c,-)

Therefore, based on the exposure dose, we divided the exposed populations into different classes. Moreover, we incorporated multiple infectious stages also by exposure level to capture disease progression. We analyzed the shedding rate of infected newts to determine the transition time from one I class to another. Each exposure threshold is determined by the variation of the shedding rate at a specific dose level. Thresholds are calculated by averaging the zoospore shed by the exposed individuals in the first recorded swab after initial exposure. We fitted experimental data (Carter et al., 2021) to estimate adult shedding rates across exposure levels (Figure 3). We assumed a quadratic function to describe the load:

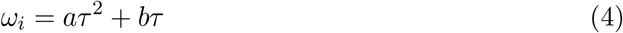

where *τ* is the number of days after the initial exposure, *a* and *b* are co-efficients. The dynamics of transitioning from one infected class to another are described below.

**Figure 3.**
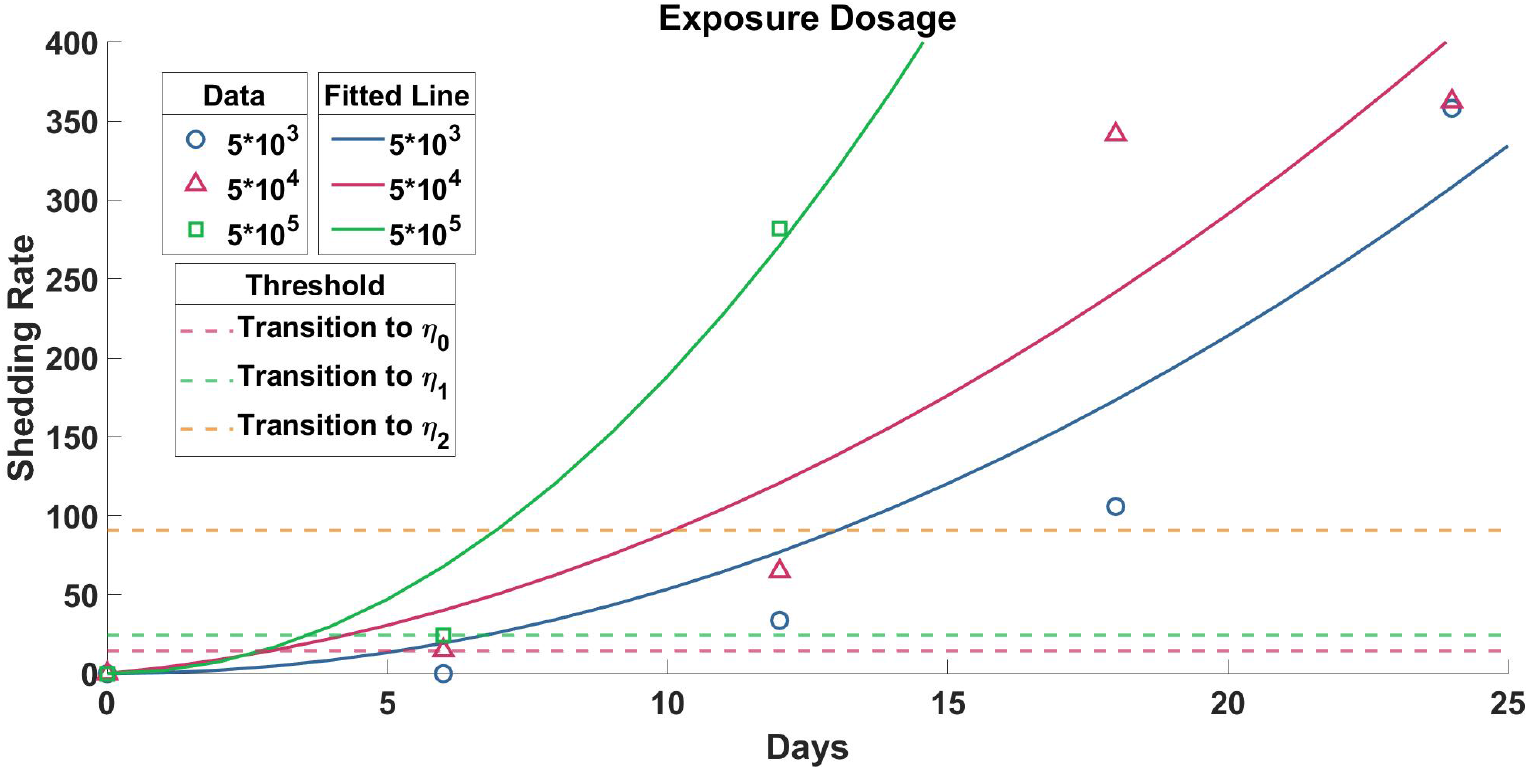
The circle, triangle, and square denote the corresponding zoospore shedding data after exposure to the zoospore concentration of 5 *×* 10^3^, 5 *×* 10^4^ and 5 *×* 10^5^, respectively. The solid blue, pink, and green lines represent the corresponding fitted lines. The dashed pink, green, and yellow lines represent the threshold for transitioning from one infectious class to another (*η*_0_, *η*_1_, and *η*_2_), depending on zoospore shedding. (see appendix Appendix A for detailed figures)

#### Disease progression and zoospore shedding rate

Based on the data from the experiment, we have subdivided our I class into four distinct compartments, *I*_0_, *I*_1_, *I*_2_ and *I*_3_ (Figure 1). Now, the susceptible population can go to any of those classes in two ways :

1. Individuals exposed to different zoospore concentrations transition directly into the corresponding *E* class, followed by the corresponding *I* class. For example, a susceptible individual exposed to zoospore concentration, 5 *×* 10^3^ will go to *E*_0_ class and follow through the *I*_0_, *I*_1_, *I*_2_ and *I*_3_. But if an individual gets exposed to 5 *×* 10^4^ concentration of zoospores, that individual will go to *E*_1_ class and eventually *I*_1_, *I*_2_ and *I*_3_ class. Due to the exposure to a higher concentration of zoospores, it will have a lower incubation period and a lower chance of survival. That is, higher exposure not only increases the probability of infection but also accelerates disease progression.
2. By direct contact with an infected individual, exposed newt begins in *E*_0_, and follows through the *I*_0_, *I*_1_, *I*_2_ and *I*_3_. This assumption reflects the brief nature of contact among eastern newts, limiting exposure to higher zoospore doses.

To estimate the transition between *I* classes, we analyzed the change in zoospore shedding rate, *ξ*, and disease progression. *Bsal* creates lesions on the skin of the infected newts. Thus, as the disease progresses, both skin zoospore load and zoospore shedding increase.

The experiment (Carter et al., 2021) provides data on zoospore-shedding rates of infected individuals exposed to a range of zoospore concentrations. When susceptible newts are directly exposed to infected newts, the zoospore shedding rate at different exposure levels correlates to distinct phases of disease progression. As seen in Table 1, the disease dynamics accelerate when susceptible newts are exposed to higher dosages. We assumed this reflects the change of shedding rate with the disease progression.

#### Data fitting and threshold

In Figure 3, we have plotted the shedding data at different exposure levels, i.e., 5 *×* 10^3^, 5 *×* 10^4^ and 5 *×* 10^5^ by using blue circles, pink triangles, and green squares respectively. We have fitted curves corresponding to these data points using Equation 4. The blue, pink, and green curves represent the fitted curve corresponding to the zoospore shedding rate of the individuals exposed at 5 *×* 10^3^, 5 *×* 10^4^, and 5 *×* 10^5^, respectively.

We calculated a threshold value from the shedding rate at different exposure levels to calculate the transition time from one infected class to another. To determine the threshold values, we considered the average amount of zoospore shed by infected individuals at the first data point. The thresholds for transitioning from *I*_0_ to *I*_1_, *I*_1_ to *I*_2_, and *I*_2_ to *I*_3_ are represented by the dashed pink, green, and yellow lines, respectively.

#### Transitioning rate (*η*_0_, *η*_1_ and *η*_2_)

We considered the shedding rates to determine the transition rate (*η*_0_, *η*_1_ and *η*_2_) of *I* classes (Figure 1). For example, from Figure 3, first, let us consider the blue curve. This blue curve indicates the exposure at 5 *×* 10^3^, and the infected individual goes through all the I classes. So, to transition from *I*_0_ to *I*_1_ class, the shedding rate needs to cross the pink dashed line. Now, the transition time from *I*_0_ to *I*_1_ is the time to cross the pink dashed line. Likewise, the transition time from *I*_1_ to *I*_2_ is the period from when the blue curve intersects the pink dashed line until it reaches the green dashed line. Similarly, the transition time from *I*_2_ to *I*_3_ is the period from when the blue curve intersects the green dashed line until it reaches the yellow dashed line.

The pink curve represents infected individuals exposed at 5 *×* 10^4^ and start at *I*_1_ instead of *I*_0_. The transitioning time from *I*_1_ to *I*_2_ is calculated as the duration from the start of the pink curve to where it intersects the green dashed line. The transitioning time from *I*_2_ to *I*_3_ is the period when the pink curve intersects the green dashed line until it reaches the yellow dashed line. In the same way, the green curve indicates that the individuals exposed at 5 *×* 10^5^, and start from the *I*_2_. The transition time for the *I*_2_ to *I*_3_ is calculated as duration from the start of the green curve until it reaches the yellow dashed line.

To calculate the transition rate from *I*_0_ to *I*_1_, *η*_0_, we used the data from individuals exposed to 5 *×* 10^3^, the blue curve. To calculate the transition rate from *I*_1_ to *I*_2_, *η*_1_, we get two estimates from the data. One estimate from the duration it takes the blue curve to transition from *I*_1_ to *I*_2_ and the second from the duration of the start of the pink curve to the *I*_2_ transition. We averaged these two estimates to parameterize *η*_1_. For the transition rate from *I*_2_ to *I*_3_, *eta*_3_, we get three estimates from the data. Two estimates from when the blue and pink curves transition from *I*_2_ from *I*_3_, and the third the green curve from its starting point to *I*_3_.

#### Probability of Transmission

The probability of disease transmission upon contact varies with the zoospore on the skin. We averaged the transmission probability upon contact based on zoospore infection load present on infected individuals Carter et al. (2021); Chowdhury et al. (2025). Transmission probability changes with the progression of diseases and exposure doses. However, for the direct transmission of disease, it does not fast-track the disease dynamics like environmental transmission. We assumed that as newts touch each other briefly, there is an increased transmission probability but no additional effects. Hence, unlike environmental transmission, all newts exposed by contact will go to the first exposed class, *E*_0_ and the corresponding *I*_0_ class.

## 5. Numerical Explorations

### 5.1. Model Simulation

Our study conducted a comprehensive simulation of the ODE system for one year with the change of initial densities of the adults (2, 4, 8, and 16 per square meter). At each initial density, the newt population experiences a rapid decline for any instance of exposure to *Bsal* (Figure 4, 4a). This occurrence highlights the significant impact of the disease, which limits the population’s ability to grow and expand beyond its initial density.

**Figure 4.**
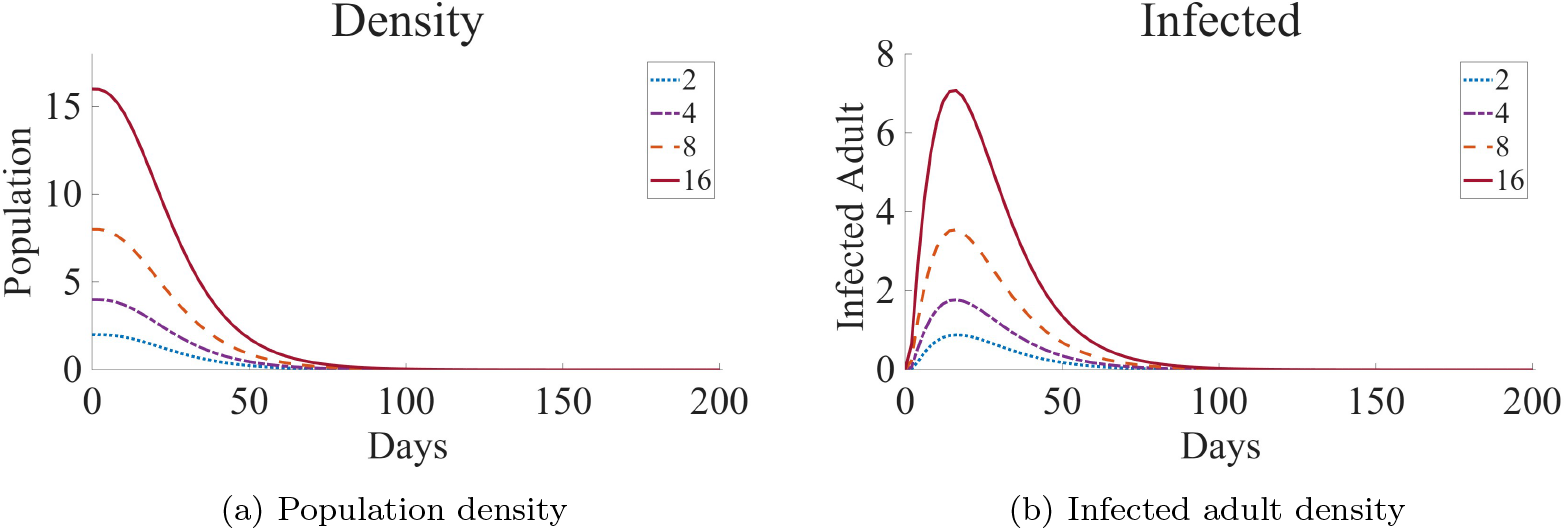
Population dynamics (a) and the number of infected newts (b) for varying initial newt population densities, 2, 4, 8, and 16 newts/m^2^.

Density-dependent simulations indicate that a lower density of 2 individuals per square meter resulted in a slower spread of the disease than a higher density(Figure 4, 4b). All the susceptible newts gets infected by *Bsal* and die within a year of the *Bsal* emergence. Moreover, we found that reduced contact at lower host density led to adult population extinction within the same timeframe as under high-density. To be more precise, we found (in Figure 5) that zoospores persist for a long time in the environment even though there is almost no susceptible newt. The persistence of zoospores in the environment prevents the recovery of the affected newt population.

**Figure 5.**
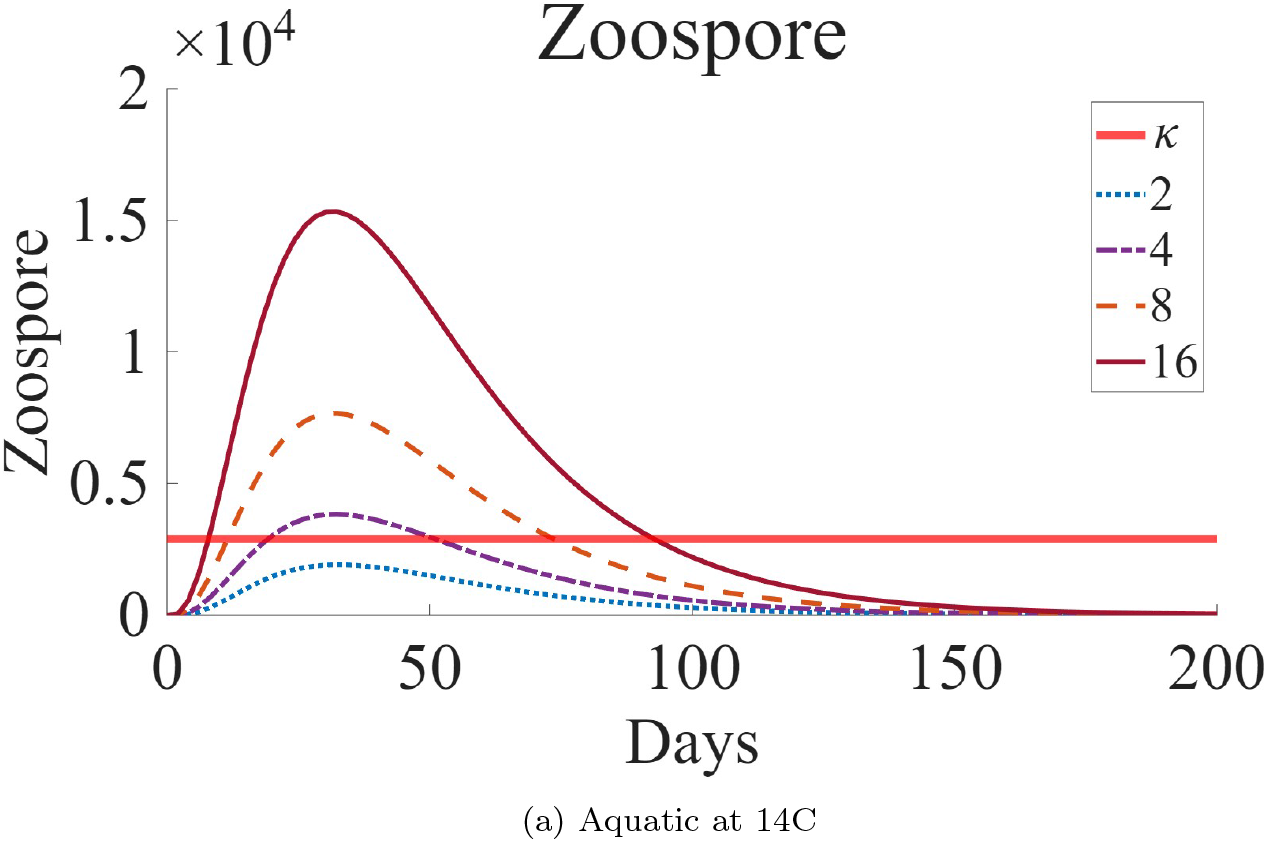
Environmental zoospore concentration (zoospore/mL) for varying initial newt population densities, 2, 4, 8, and 16 newts/m^2^.

## 6. Basic Reproduction Number

The basic reproduction number, or *ℛ*_0_, estimates the transmission rate of infectious diseases (Dietz, 1993; Delamater et al., 2019; Heffernan et al., 2005). It indicates the average number of secondary infections that result from one infected individual in a susceptible population. The disease will die down if *ℛ*_0_ *<* 1. On the other hand, if *ℛ*_0_ *>* 1, then the disease will spread and can become an epidemic.

Using the next-generation matrix approach (Roberts and Heesterbeek, 2013; Van den Driessche, 2017; Diekmann et al., 2010), we have calculated the basic reproduction number. The analytical representation of the basic reproduction number of the system is below, and the detailed calculation is shown in the Appendix C.

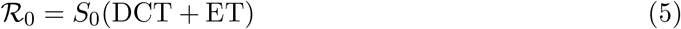

where DCT stands for direct contact transmission, and ET stands for environmental transmission due to the zoospore pool in the environment. The expression for DCT and ET is,

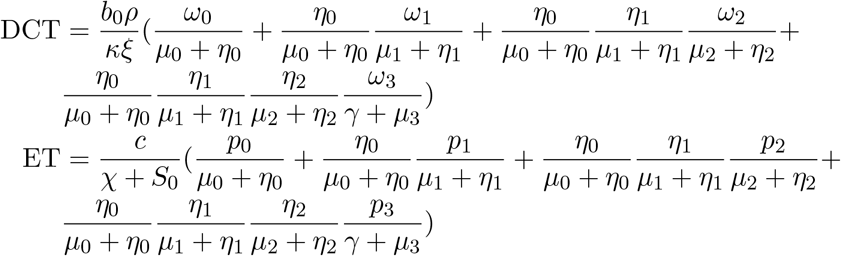

### 6.1. Parameter Sensitivity Analysis

We analysed the ℛ_0_ expressions using Latin Hypercube Sampling (LHS) and Partial Rank Correlation Coefficient (PRCC) method. (Equation 6) (Figure 6). The LHS/PRCC technique, developed by Gomero (2012), explores the multi-dimensional parameter space of the complete model globally. The PRCC values for adults are presented in Figure 6, and the monotonicity of the parameters is in the Appendix D.

**Figure 6.**
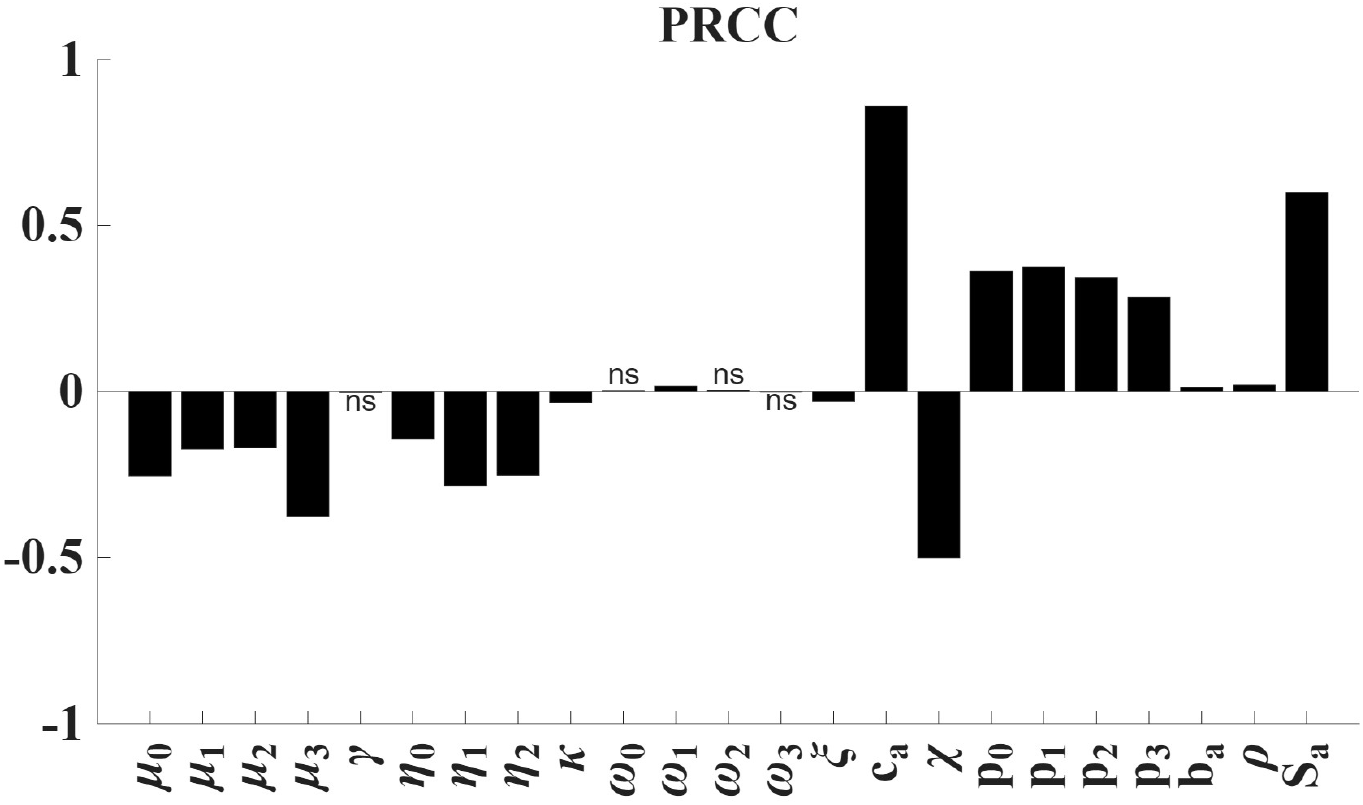
Partial rank correlation coefficient (PRCC) for each parameter in the Latin hypercube sampling (LHS) for ℛ_0_. Values marked ns are non-significant (*P >* 0.05).

Figure 6 demonstrates parameters significantly influencing the basic reproduction number. There are two types of correlations: positive and negative. Positively correlated parameters raise ℛ_0_ with an increase in parameter value, while negatively correlated parameters do the opposite. ℛ_0_ has a strongly negative connection with the disease-induced death rates (*µ*_0_,*µ*_1_,*µ*_2_,*µ*_3_) and the zoospore degradation rate *ξ*. However, a positive correlation exists between the contact parameter *c*_*a*_ and the transmission probability of disease following contact. All the other positively linked metrics are dwarfed by the high contact rate among adults.

### 6.2. Numerical Explorations of ℛ_0_

We simulated the ℛ_0_ values by varying direct contact rates, zoospore degradation rate,*ξ*, and initial population density, *S*_0_. Lower zoospore degradation increases the zoospore pool, raising contact and transmission likelihood. Therefore, we paired the zoospore degradation rate with the direct contact rate. On the other hand, we paired the population density with the direct contact rate to explore the impact of density on ℛ_0_. From the Figure 7, it is evident that ℛ_0_ increases sharply with higher direct contact and lower zoospore degradation rate *ξ*. However, high contact rates overshadow the contribution of environmental transmission. Conversely, lower host density and reduced contact lead to a decline in ℛ_0_.

**Figure 7.**
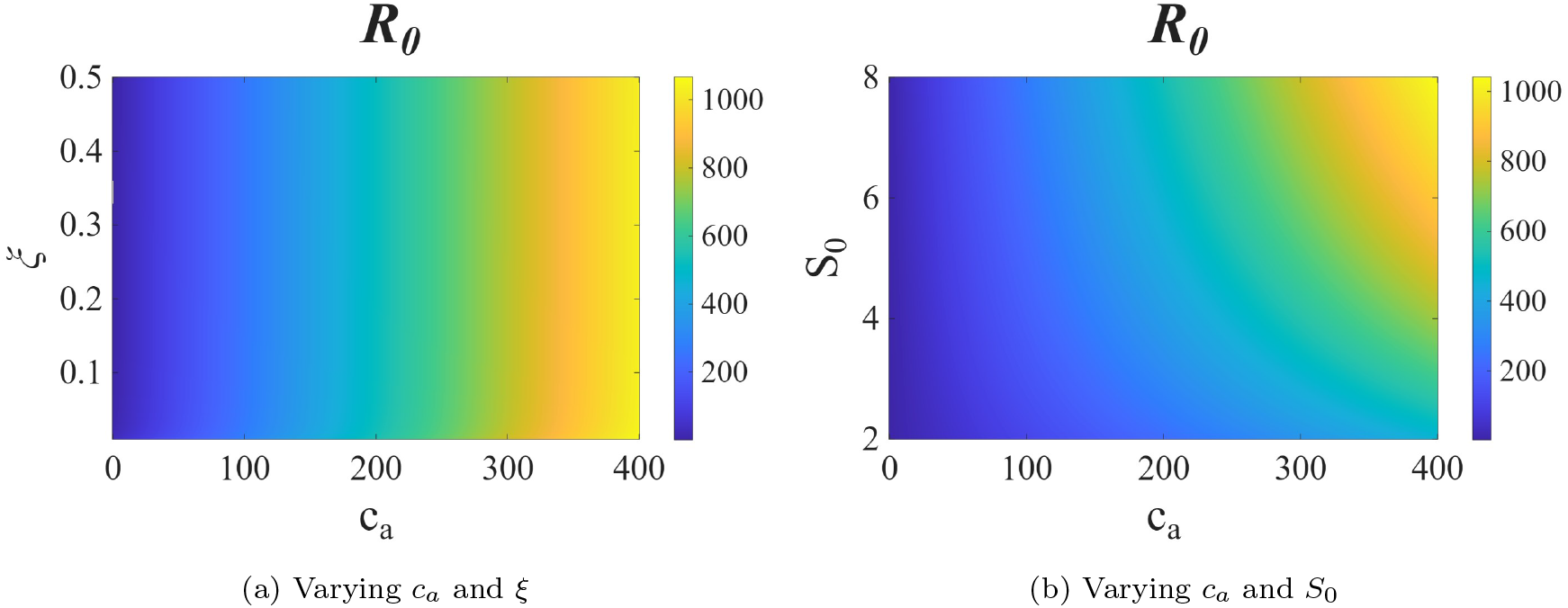
Simulation of ℛ_0_ on varying contact rate, *c*_*a*_ and zoospore degradation rate, *ξ* (7a) and contact rate, *c*_*a*_ and initial population density, *S*_0_ (7b)

### 6.3. Dominant Transmission Pathway

Adult newts become infected *Bsal* in two pathways: direct contact with the infected newt and indirect exposure to environmental zoospores. From the experiments, we have seen that adults have a high direct contact rate. In contrast, environmental transmission depends on zoospore survival and the likelihood of newt contact with them. Accounting for these uncertainties, we explored the dominance of each transmission pathway by varying the direct contact rate (Figure 8,8a) and the zoospore degradation rate (Figure 8,8b). Simulations suggest that without a high contact rate, the disease transmission will be dominated by the environmental pathway (Figure 8).

**Figure 8.**
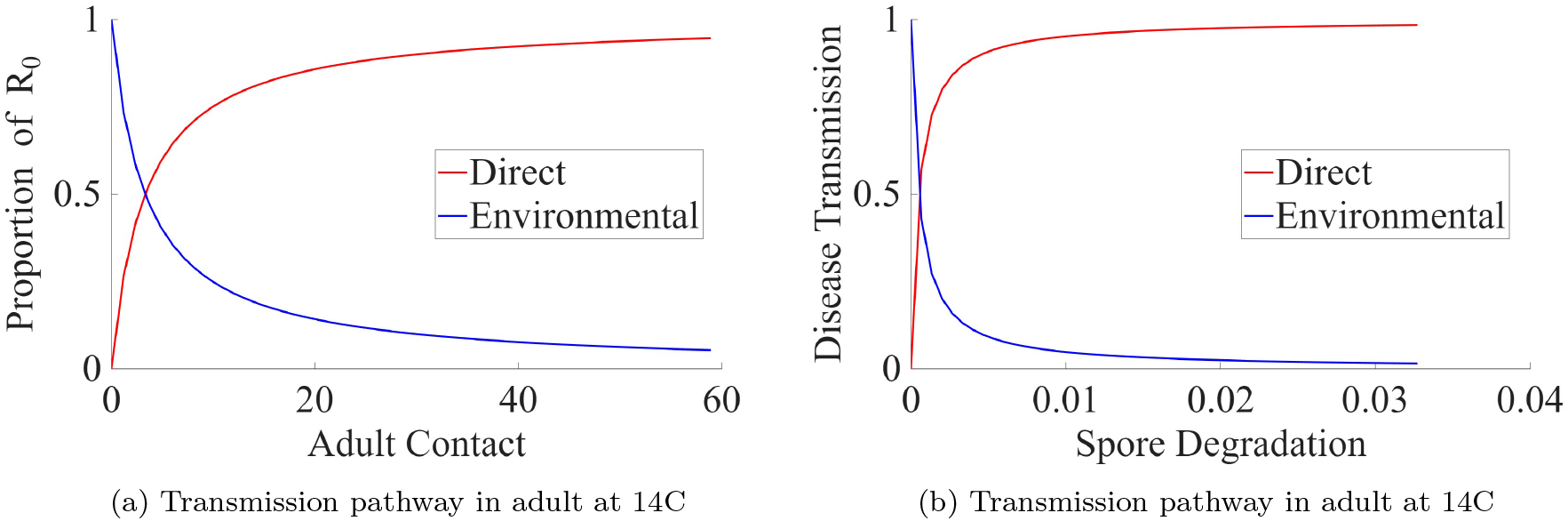
Dominant transmission pathway in the aquatic environment for adults with varying direct contact rate and zoospore degradation rate.

## 7. Micro-predators and Spill Over of *Bsal*

In a recent paper by Gray et al. (2023), a wide range of amphibian species have been found susceptible to *Bsal*, with some capable of acting as a disease carrier. They evaluated *Bsal* invasion risk through dose–response experiments on 35 North American amphibian species, observing 74% infection and 35% mortality, with both salamanders and frogs developing chytridiomycosis. The ID50 ranged from 2897 to 52891331 across species, which included four very high–risk, two high–risk, and three moderate–risk species For this, minimizing environmental zoospore concentration is essential to prevent *Bsal* spillover to other species and habitats. Specifically, reducing environmental zoospore concentration can support the survival of species that are highly susceptible to *Bsal* and have a low ID50. For this, we have extended our environmental load-driven transmission model to include the consumption of zoospore by micro-predator. *Daphnia* is a natural predator of zoospores in the water. We conducted experiments at the University of Tennessee at Knoxville on *Bsal* consumption of *Daphnia magna* following procedures described in Appendix E and detailed in Appendix E. From this experiment, we quantified the zoospore consumption by *Daphnia magna* and analyzed the impact of zoospore consumption on environmental transmission. With this, our environmental load-driven transmission model structure remain same except for a change in the zoospore degradation rate. Alongside *ξ*, another parameter *α*, represents the consumption of zoospores by *Daphnia* within the *Z* compartment. With this addition, the environmental zoospore tracking equation Equation 3 becomes -

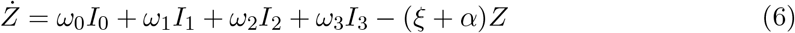

To examine how changes in zoospore dynamics influence environmental transmission, we simulated the model under three severity levels: low, medium and high (Table 3) with and without micropredator. We varied zoospore shedding (*ω*), infectious stage duration (*η*), and zoospore degradation in the first infectious stage (*µ*_*aa*_). These parameters directly regulate zoospore concentration and persistence in the environment.

**Table 2.** Parameter Table.

| Parameter | Meaning | Values | Source |
| --- | --- | --- | --- |
| $\eta_i, i = 0, 1, 2$ | Transitioning from one $I_i$ to $I_{i+1}$ | day<sup>-1</sup> | From Unpublished data (Carter et al., 2021) |
| $\sigma_i, i = 0, 1, 2, 3$ | Latency period | Table 1 day<sup>-1</sup> | Estimated from the experiment of (Carter et al., 2021) |
| $\kappa$ | ID50 of adult | 2895 zoospores/ml | (Carter et al., 2021) |
| $\gamma$ | Recovery rate of adult | 0 day<sup>-1</sup> | Assumed |
| $\mu_i, i = 0, 1, 2, 3$ | Disease induced death rate | Table 1 day<sup>-1</sup> | Estimated from the experiment of (Carter et al., 2021) |
| $\rho$ | Percentage of zoospores that encysted | 75% | (Islam et al., 2021) |
| $\xi$ | Zoospore degradation rate | 1/25 day<sup>-1</sup> | (Stegen et al., 2017) |
| $\omega_i, i = 0, 1, 2, 3$ | Zoospore shedding rate | Table 1 day<sup>-1</sup> | From the experiment in (Carter et al., 2021; Chowdhury et al., 2025) |
| $P_i, i = 0, 1, 2, 3$ | Prob. of transmission given contact between adults | Table 1 | From the experiment in (Chowdhury et al., 2025) |
| $b_a$ | Contact between zoospore and adult | Assumed | 0.1 (Islam et al., 2021) |
| $\alpha$ | Zoospore Consumption Rate by Daphnia per day | 0.0567 | Appendix E (Gray and Carter, 2022-2024a) |

**Table 3.** Parameter sets for low, medium, and high severity scenarios with and without micro-predators.

| Parameter | Low | Medium | High |
| --- | --- | --- | --- |
| $\eta$ (infectious staying period) | 1/5.233 | 1/20.233 | 1/40.233 |
| $\omega$ (zoospore shedding) | 142.82 | $142.82 \times 5$ | $142.82 \times 10$ |
| $\mu_{aa}$ (first-stage degradation) | 1/30.6 | $1/(2 \times 30.6)$ | $1/(3 \times 30.6)$ |

It is evident from Figure 9 that *Dapnia* effectively reduces the zoospore concentration in the environment. Considering this, we simulated the model to assess whether the presence of a micropredator significantly alters environmental transmission dynamics. Figure 10 shows a reduction in environmental transmission corresponding to changes in zoospore concentration. To explore the broader impact, we simulated the model under reduced environmental transmission. Despite the decline in zoospore levels due to micropredation, no significant improvement in adult survival was observed. This is likely due to the high direct contact rate among adults. Thus, while micropredators may not affect *Bsal* transmission within eastern newts, they could still play a key role in preventing spillover.

**Figure 9.**
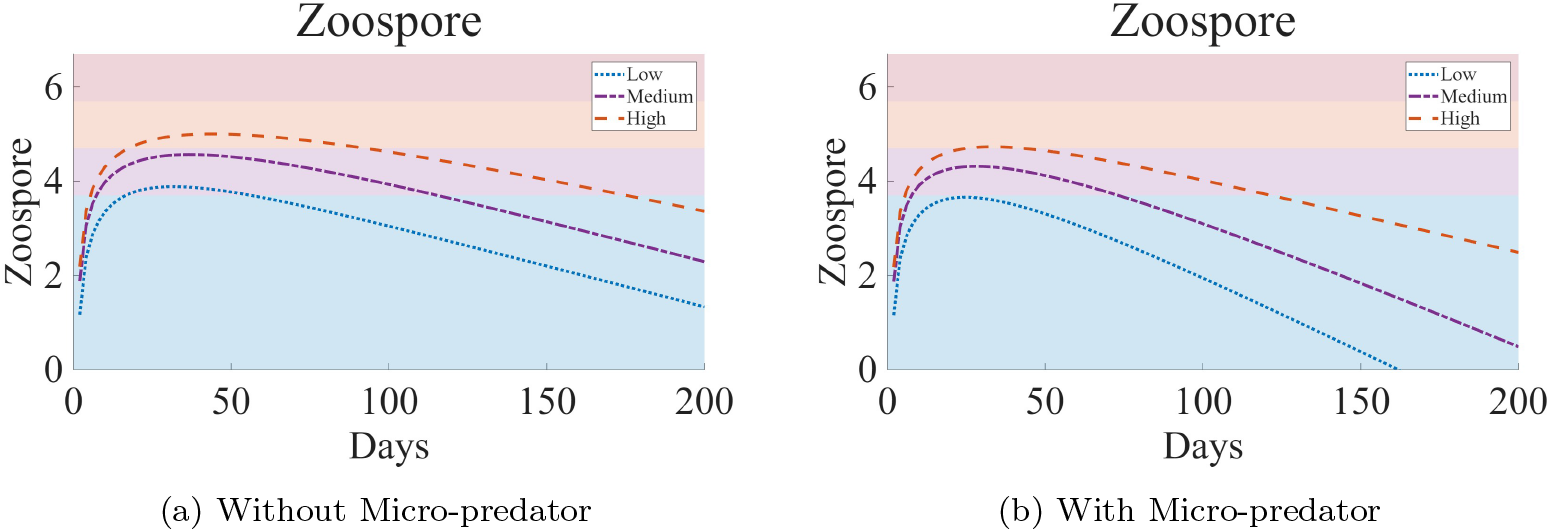
Zoospore concentrations over time without (left) and with (right) micro-predators for scenarios of low (blue dotted), medium (violet dotted-dashed), and high (orange dashed) severity scenarios as defined in Table 3. The horizontal shaded bands indicate the log transformed zoospore thresholds corresponding with environmental transmission in Equation (2): 0 to 5*×*10^3^ (blue), 5*×*10^3^ to 5*×*10^4^ (purple), 5*×*10^4^ to 5*×*10^5^ (orange), and 5 *×* 10 to 5 *×* 10 (pink).).

**Figure 10.**
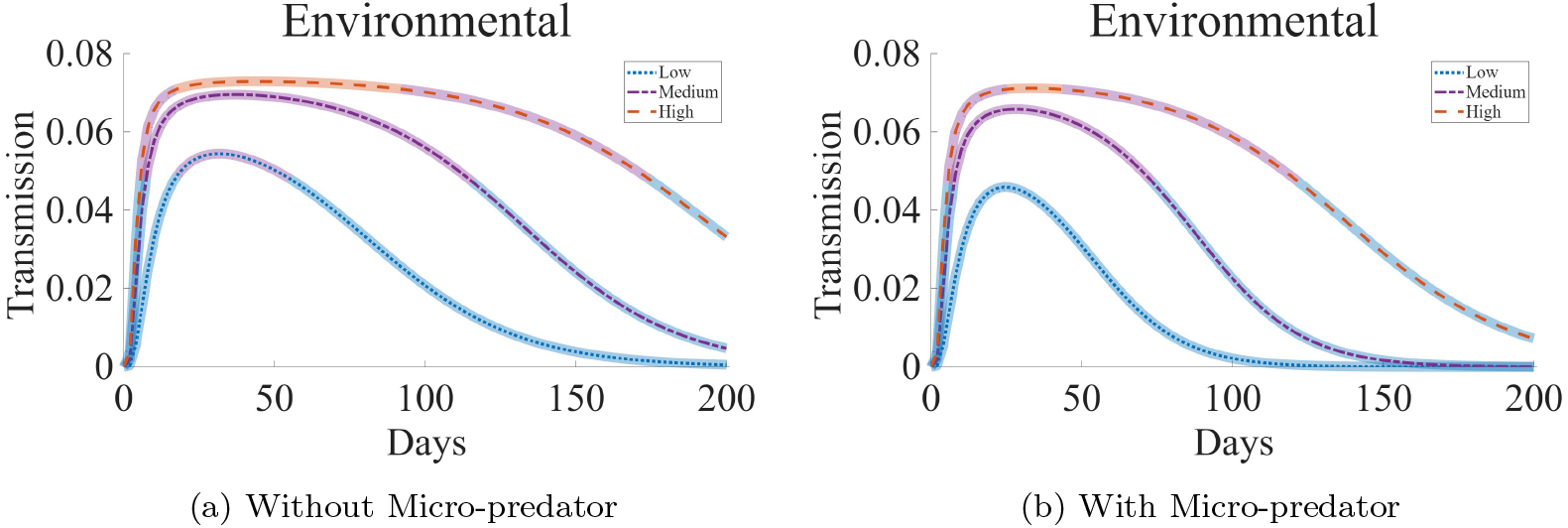
Environmental transmission among adults without (left) and with (right) micro-predator for scenarios of low (blue dotted), medium (violet dotted-dashed), and high (orange dashed) severity scenarios as defined in Table 3. The horizontal shaded regions around the curves indicate the log transformed zoospore thresholds corresponding with environmental transmission in Equation (2): 0 to 5*×*10^3^ (blue), 5*×*10^3^ to 5*×* 10^4^ (purple), and 5 *×* 10^4^ to 5 *×* 10^5^ (orange), corresponding to the shaded regions in Figure 9.)

## Discussion

This study introduces a novel compartmental modeling framework that captures exposure-dependent disease dynamics with multiple transmission pathways. Traditional SEIR-type models often assume uniform infectiousness and constant transmission probability. Our model incorporates heterogeneity in exposure dose, progression, and mortality by structuring exposed and infectious compartments based on pathogen load. This enables a more realistic representation of diseases, where infection severity and outcome are dosedependent. Such dose-dependent dynamics are common in fungal, viral, and bacterial infections that involves environmental transmission. We also extended the model by incorporating a predator-zoospore interaction module. Micropredators can reduce the likelihood of pathogen introduction and zoospore transmission between heterospecifics during outbreak events.

We applied our model to address the serious ecological threat posed by the chytridiomycosis causing fungal pathogen *Bsal*. Our simulations unfold interconnected patterns of *Bsal* transmission within susceptible, infected, and zoospore pools in the environment. The model captures patterns of disease progression and outbreak severity in varying ecological scenarios. Simulations indicate that both the initial exposure dose and host density affect outbreaks and host mortality. The model also captures dynamic feedback between hosts and the environmental zoospore reservoir by explicitly modeling host-zoospore dynamics. One key strength of the model is its ability to identify dominant transmission routes under different ecological conditions. When direct contact rates are high, transmission occurs mainly through host-to-host interaction. When contact is limited, environmental transmission becomes the dominant pathway. This is especially true when zoospore decay is slow. These findings highlight the need to account for both direct and environmental components when modeling pathogens with multiple transmission pathways.

Model’s sensitivity analysis further highlights the utility of exposure-based stratification. The basic reproduction number (ℛ_0_) was most sensitive to the direct contact rate, initial host density, and environmental zoospore degradation rate. This aligns with ecological intuition, but also shows the model’s capacity to quantify which factors drive outbreak potential. Even though our basic reproduction number in the adults is quite high but this has been seen in other amphibian diseases (Edelstein-Keshet, 2005; Wilber et al., 2017; Peace et al., 2019). The estimates of our ℛ_0_ value is similar to pathogens spread by mosquitoes at high densities. (Davidson, 1955; Davidson and Draper, 1953). Therefore, even if our ℛ_0_ estimation is biased by several order of magnitude, *Bsal* represents a severe threat to caudate species around the globe. Importantly, our framework allows for adjusting these estimates under varying environmental and behavioral scenarios, supporting flexible disease forecasting. To further demonstrate the model’s extensibility, we incorporated a predator-zoospore interaction module. This module represents zoospore consumption by aquatic micro-predators. Predation reduced zoospore concentrations but had little effect on disease progression as contact rates were high. However, predation substantially decreased the spill-over risk, pointing to a possible ecological intervention strategy. This extension demonstrates that our model can incorporate biotic interactions, such as the effects of predators on zoospore levels and transmission framework.

In summary, our model bridges the gap between dose-dependent transmission biology and classical compartmental modeling. The framework is adaptable to host–pathogen systems where pathogen load, environmental persistence, and ecological feedbacks are important. Future research should explore spatial extensions of the model. Furthermore, future models should explore spillover events to evaluate transmission dynamics within complex amphibian communities that have mixed levels of susceptibility. These models could then help explain patterns found in pathogen surveillance studies.

## Data Availability

Data will be available upon reasonable request.

## 9. Acknowledgement

This work was supported by the National Science Foundation grant DEB-1814520. Also, MJG was partially supported by USDA National Institute of Food and Agriculture Hatch Project-7001607.

## Supplementary Material

### A. Transitioning rate, *η*_**0**_, ***η***_**1**_ **and *η***_**3**_

### B. Basic Reproduction Number, *ℛ* _**0**_

For disease dynamics, we have considered a SEIR-type (Susceptible, Exposed, Infected, and Recovered) model with multiple exposed (E) and infected (I) compartments for the adult population. Depending on the disease progression and the zoospore amount on the skin, we have divided the infected class into four compartments. We did not consider the four exposed classes for the calculation of the basic reproduction number as the exposed class depends on the change in incubation periods which depends on the zoospore concentration in the environment. As the definition of the basic reproduction number suggests, the average number of cases of an infectious disease arising by transmission from a single infected individual, in a population that has not previously encountered the disease, the amount of the zoospore in the environment never exceeds the threshold zoospore amount. As the zoospore amount in the environment depends directly on the shedding of the infected specimen, therefore the amount of zoospore amount will never be able to pass the initial threshold. For this, we have considered only one exposed class with four infected class.

To calculate the basic reproduction number, we have utilized the next-generation matrix approach. By definition of basic reproduction number, we have set the equations to zero and solved to find the disease-free equilibrium. The disease-free equilibrium of the model is *X*_0_ = (*S*_*a*_, 0, 0, 0, 0, 0, 0). By reorganizing the system (1), we have considered the exposed and infected classes for the Jacobian matrix *J* . Evaluating the Jacobian matrix at *X*_0_,

### C. Basic Reproduction Number, ℛ _**0**_

We have utilized the next-generation matrix approach to calculate the basic reproduction number. By definition of basic reproduction number, we have set the equations to zero and solved to find the disease-free equilibrium. The disease-free equilibrium of the model is *X*_0_ = (*S*_0_, 0, 0, 0, 0, 0, 0). By reorganizing the system (1), we have considered the exposed and infected classes for the Jacobian matrix *J* . Evaluating the Jacobian matrix at *X*_0_,

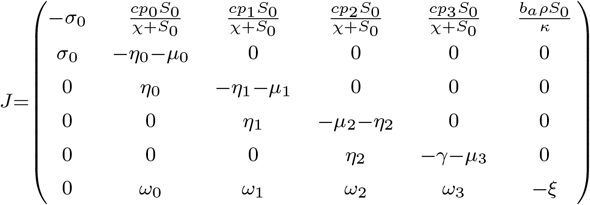

The linearized system changes according to the following set of equations for minor perturbations *z* = (*S*_0_, *E*_0_, *I*_0_, *I*_1_, *I*_2_, *I*_3_, *R*_0_, *Z*) near the *X*_0_

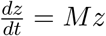

Here,

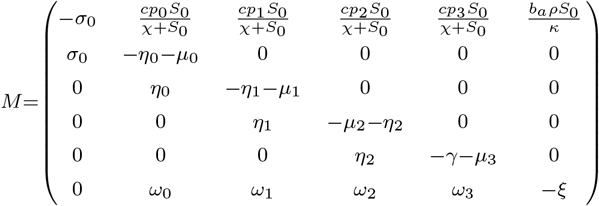

We decompose the matrix M into transmission (T) and transition (Σ) matrices respectively, obtaining

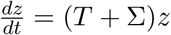

Where

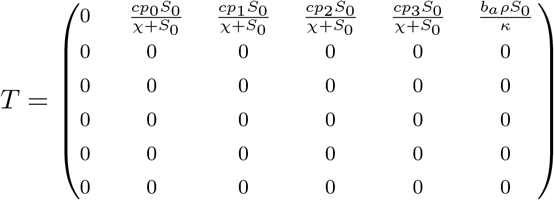

And

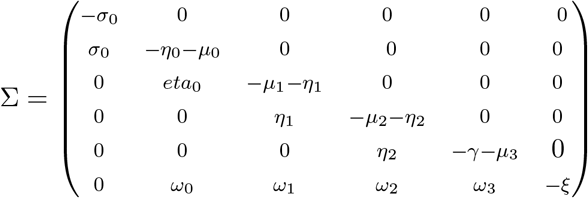

Now the non-zero eigenvalues of ™*T* Σ^™1^ are,

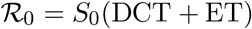

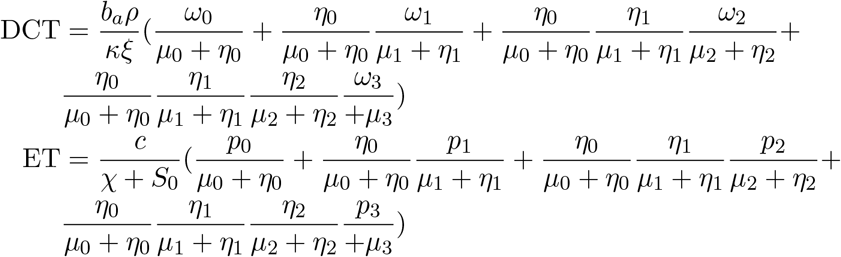

### D. Monotonocity

All parameters exhibit a monotonic relationship with output measures Figure 2.

All parameters exhibit a monotonic relationship with output measures Figure 2.

### E. Zoospore consumption by aquatic micropredators

We conducted laboratory experiments to quantify the consumption of *Batrachochytrium salamandrivorans (Bsal)* zoospores by aquatic micropredators. *Daphnia magna* was used as the model zooplankton species and obtained from Carolina Biological. Three density treatments were prepared (15, 30, and 60 individuals mL^™1^). One milliliter of each *D. magna* suspension was added to wells of a 24-well flat-bottom microplate. Control wells contained the same water source without zooplankton.

*Bsal* zoospores (isolate AMFP 13/1), originally isolated from a morbid fire salamander (*Salamandra salamandra*; (Martel et al., 2013)), were cultured at the University of Tennessee. Zoospores were harvested and enumerated following established methods ((Carter et al., 2020)). Zoospores were diluted to 2 *×* 10^6^ zoospores mL^™1^. One milliliter of the zoospore stock was added to each well, yielding a final concentration of 1 *×* 10^6^ zoospores mL^™1^.

Three independent trials were conducted. Each trial included all *D. magna* density treatments and control wells, with six replicates per treatment. Zoospore concentrations were quantified at three and six hours post co-incubation. Ten microliters from each well were enumerated using disposable improved Neubauer hemocytometers. Wells were gently mixed prior to sampling. Zoospore consumption rates (zoospores mL^™1^ h^™1^) were calculated as the difference between control wells and *D. magna* treatments. We quantified zoospore consumption by *Daphnia magna* as the number of *Batrachochytrium salamandrivorans* zoospores removed per mL per day. Rates were normalized by the initial zoospore concentration (10 zoospores mL^1^) and averaged across predator densities and sampling times to obtain an effective per-capita clearance rate.

### F. Severity Table

**Figure 1.**
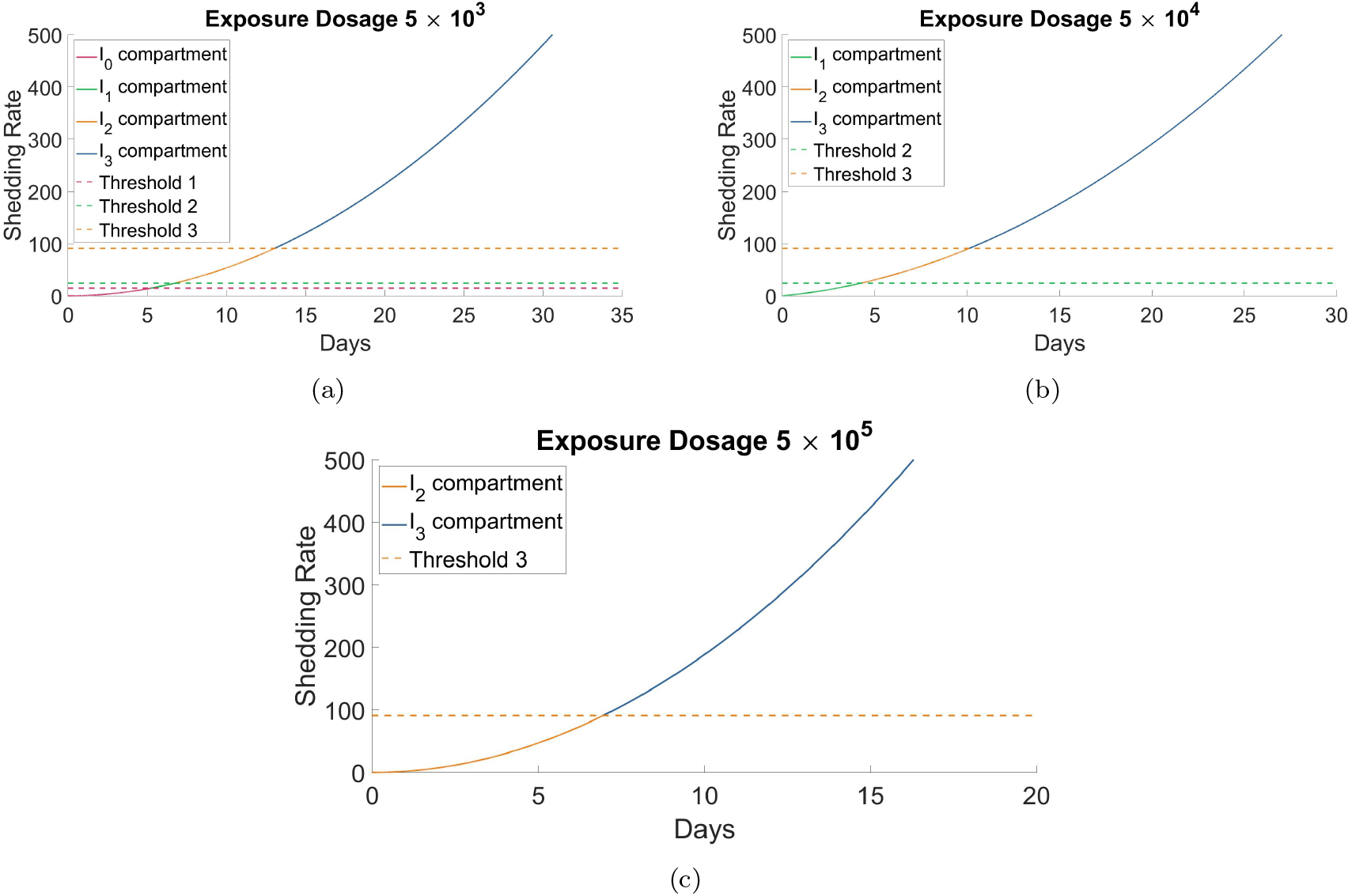
The curve’s colors represent segments of different compartments: pink for *I*_0_, green for *I*_1_, yellow for *I*_2_, and blue for *I*_3_. Figures (a), (b), and (c) illustrate transitions between infection classes *I*_0_, *I*_1_, *I*_2_, and *I*_3_ at varying initial exposures: (a) 5 *×* 10^3^, (b) 5 *×* 10^4^, and (c) 5 *×* 10^5^. The curves in (a), (b), and (c) are represented by blue, pink, and green curves in Figure 3.

**Figure 2.**
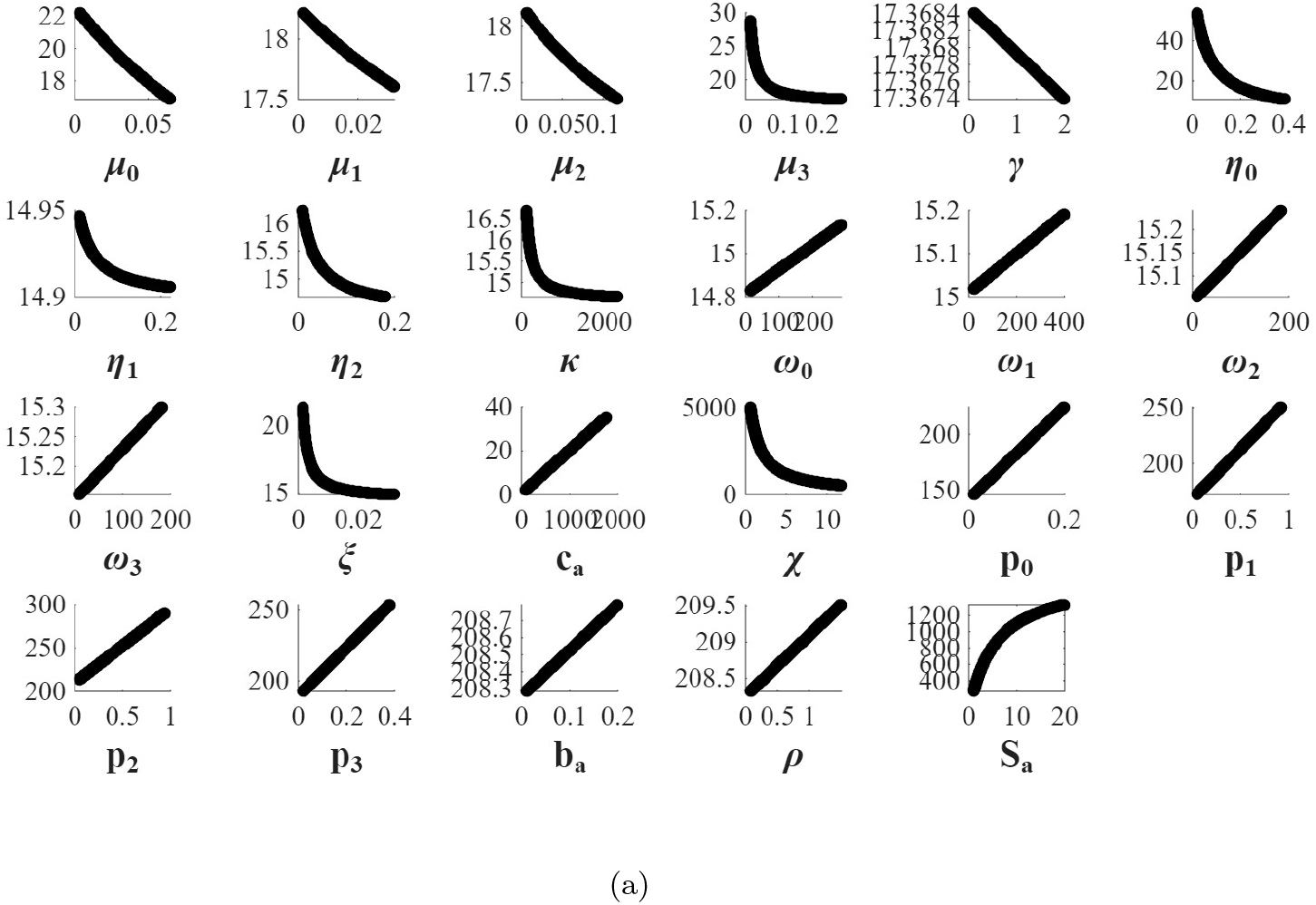
Monotocity

